# Detection of SARS-CoV-2 in Schools Using Built Environment Testing in Ottawa, Canada: A Multi-Facility Prospective Surveillance Study

**DOI:** 10.1101/2023.03.03.23286750

**Authors:** Nisha Thampi, Tasha Burhunduli, Jamie Strain, Ashley Raudanskis, Jason A. Moggridge, Aaron Hinz, Evgueni Doukhanine, Castellani, Fralick, Rees Kassen, Janine McCready, Caroline Nott, Wong, Derek R. MacFadden

## Abstract

Classroom and staffroom floor swabs across six elementary schools in Ottawa, Canada were tested for SARS-CoV-2. Schools in neighbourhoods with historically elevated COVID-19 burden had lower environmental swab positivity. Environmental test positivity did not correlate with student grade groups, school-level absenteeism, pediatric COVID-19-related hospitalizations, or community SARS-CoV-2 wastewater levels.

**Summary:** Environmental SARS-CoV-2 sampling was performed in six schools in Ottawa, Canada. The percentage of floor swabs detecting SARS-CoV2 was not correlated with absenteeism, pediatric hospitalizations, or wastewater data. Schools in neighbourhoods with previously elevated COVID-19 rates had lower test positivity.

## Background

Environmental surveillance for SARS-CoV-2 has been an effective public health tool for monitoring COVID-19 in the population, with the most studied approach being wastewater sampling. (1) Surface sampling within the built environment may be a more spatially refined approach for surveillance in specific contexts. For example, a recent multi-centre study across 10 long-term care homes demonstrated that the percentage of positive swabs for SARS-CoV-2 correlates with cases and outbreaks of COVID-19.(2) A strong correlation between environmental sampling and cases of COVID-19 has also been shown in the hospital setting.(3,4) While SARS-CoV-2 has been detected in non-healthcare settings such as schools, it is unclear whether trends correlate with community prevalence.(5–7) We sought to determine the relationship between SARS-CoV-2 swab positivity in the school environment and community indicators of COVID-19 prevalence.

## Methods

A prospective surveillance study was conducted in six publicly funded elementary schools over a 12-week period, from March 28 to Jun 17, 2022, in Ottawa, Ontario. Floors of every classroom, gymnasium and staff lounge in each school were sampled twice weekly by research or school staff trained by the research team, with a minimum two-day interval between most collection times. Sites identified by the research team and school administration were in high-traffic areas, including entrances, vestibules, and handwashing stations in the room. The same 2-by-2-inch areas of the floors were swabbed each time, with one sample collected from each room and processed by the laboratory within the week of collection using previously validated protocols.(2,3) Wastewater data were obtained from publicly available sources; weekly means and 95% confidence intervals (CIs) were calculated from daily N1 and N2 target values, with the quantity of SARS-CoV-2 RNA expressed as a proportion relative to Pepper mild mottle virus (PMMoV) RNA.(8,9) Permission was received from boards and principals prior to each school visit, and school communities were notified by the principal prior to starting sample collection.

COVID-19 vaccine eligibility had been extended to children ages 5 to 11 years in November 2021. Ontario lifted mandatory masking policies in schools and other indoor settings apart from public transit on March 21, 2022, when students were set to return to in-person learning following Spring Break. School-based mask recommendations shifted from mandatory to strongly recommended. The largest Ottawa school board reintroduced mandatory masking between April 13 and May 30 in response to high community prevalence of SARS-CoV-2. In Ontario, students enroll in Grade 1 in the year of their sixth birthday. Thus, the youngest child in Junior Kindergarten may be 3 years old, and the oldest child in Grade 8 may be 14 years old.

The main outcome was SARS-CoV-2 detection on floor surfaces. Data were analyzed at the level of the student classroom, staffrooms and school, and compared to publicly available data on historical SARS-CoV-2 burden in Ottawa neighbourhoods served by these schools, as well as school-level student and staff absenteeism rates, city-level wastewater signals, and number of COVID-19-associated pediatric hospitalizations at CHEO, the sole pediatric acute care facility in the region, which was submitted daily to the Ontario Ministry of Health.(8–10) During this period, testing was restricted to children admitted with infectious symptoms. Ethics review was waived by the Research Ethics Board of the University of Ottawa as no identifiable human data were collected.

Statistical analyses were performing using the R programming language (v4.2.2).(11) Confidence intervals on proportions (positivity, absenteeism rates) were computed using the Wilson method. Generalized linear mixed models (GLMMs) were created using ‘glmer’ from the R package ‘lme4’ (v1.1-29).(12) Classroom-level data were grouped into Junior and Senior Kindergarten, Grades 1-3, Grades 4-6, and Grades 7-8 for analysis of age group effects. Swab positivity and absenteeism rates were grouped into semiweekly bins to compute school-level means for correlation analysis. We calculated the Spearman correlation coefficient (*r*) between the presence of SARS-CoV-2 on floors, absenteeism, pediatric hospitalizations, and levels of SARS-CoV-2 detected in community wastewater surveillance.

## Results

During the 12-week study period, 2,860 floor samples were collected across six schools. Overall test positivity for SARS-CoV-2 RNA was 20%, with rates ranging from 10% to 36% among schools (median 16.5%) and from 0% to 77.8% among rooms (median 15.8%). Weekly aggregated floor-swab positivity across schools remained between 10-30% over the study period, with small peaks observed during early-to-mid-April, early-to-mid-May, and early June (fig. 1). Four schools had mandatory masking policies in place between April 13 and May 30, 2022. We found no correlation between swab positivity and absenteeism rates within schools, irrespective of the presence of mandatory masking policies (Supplemental Figure 1).

**Figure 1.**
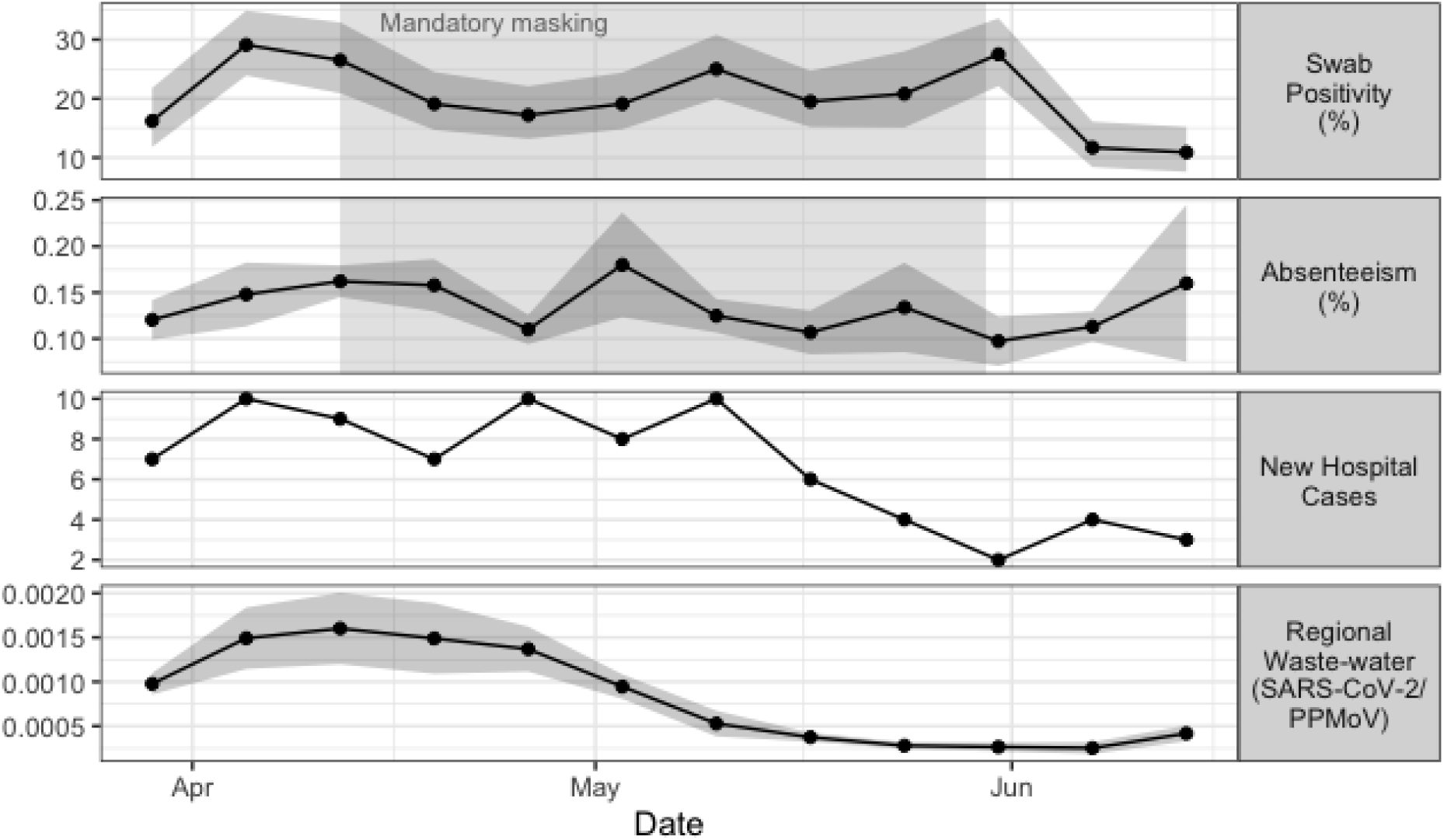
Weekly time-series. (panels top to bottom): weekly mean positivity of environmental PCR tests from swabs collected at six schools (% and 95% CI); weekly mean absenteeism at the same schools (% and 95% CI); weekly counts of new SARS-CoV-2 cases at the regional children’s hospital; and Ottawa regional waste-water SARS-CoV-2 detection relative to PPMoV (mean and 95% CI of N1 and N2 targets). Points display either means or count values, shaded ribbons around the means represent the 95% CI, where applicable. Shaded boxes indicate the period during which mandatory masking policies were in effect at four of the six schools.

There was no significant correlation between the weekly floor-swab positivity of schools and regional wastewater signal, school absenteeism rates, or number of pediatric COVID-19-related hospitalizations during the same week (Spearman’s *r* = 0.235, -0.039, and 0.285 respectively; *p* > 0.05; fig. 1). Similarly, absenteeism rates were not significantly correlated with wastewater signal or hospitalizations (Spearman’s *r* = 0.52 and 0.25, respectively; *p* > 0.05); however, the regional wastewater detection and pediatric COVID-19-related hospitalizations were strongly correlated during the study period (Spearman’s *r* = 0.75; *p* = 0.005). Overall floor-swab positivity rates at the school level were not correlated with the prevalence of COVID-19 prior to 2022 in the surrounding neighbourhood (Spearman’s *r* = -0.6, *p* = 0.24; Table 1). Floor swab positivity did not change significantly by day of the week when floors were swabbed (data not shown).

**Table 1:**
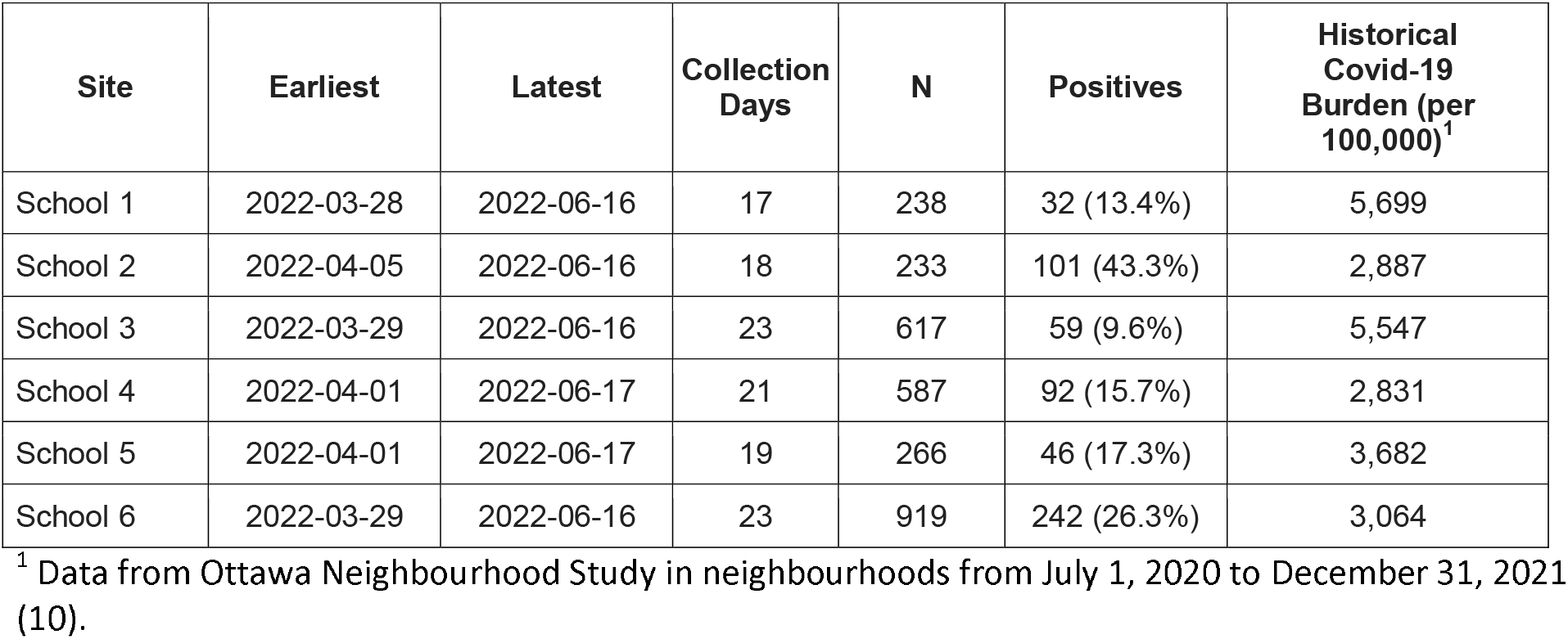
Summary of surface surveillance of SARS-CoV-2 conducted at six schools and historical case rates for the surrounding neighbourhoods.

Swab positivity was also examined at the grade level by classroom (Supplemental Figure 2). Staff rooms, followed by classrooms with children in Junior and Senior Kindergarten generally had higher swab positivity, while Grades 7-8 classrooms had lower swab positivity throughout the period of study compared to other classrooms. A likelihood ratio test of nested GLMMs failed to find a significant fixed effect of age (*p* = 0.28), suggesting that apparent differences in swab positivity rates between age groups arise from the differences in the composition of age group (*e*.*g*., roughly 50% of the grades 7-8 classrooms are situated in School #3, which has the lowest positivity among schools).

## Discussion

In our 12-week prospective study across six publicly funded elementary schools, SARS-CoV-2 was detected 20% of the time on floors of classrooms, gyms, and staff areas, and did not correlate with student and staff absenteeism or community wastewater signals at the school or aggregate level. A negative but non-significant correlation was found between test positivity in schools and prior COVID-19 infection rates in neighbourhoods which may have served as proxy for community immunity among student populations. The variability in test positivity among schools suggest hyper-localization of SARS-CoV-2 transmission during this period.

Strengths of the study include serial sampling of floors during a community surge of SARS-CoV-2, examination of the impact of school-based mandatory masking policies, and comparisons with community metrics, namely wastewater signals, historical neighbourhood COVID-19 infection rates, and pediatric COVID-19 disease requiring hospitalization.

School-based surveillance of respiratory infections is a key aspect to understanding the burden and dynamics of infection transmission through schools and workplaces, households and social networks (6,13–17). Environmental surveillance has been proposed as an additional layer to inform pandemic and endemic disease surveillance, given the limitations of human-based testing as a surveillance strategy to inform public health action.(18)

Wastewater testing has demonstrated usefulness in assessing SARS-CoV-2 levels in the community and acute care, however it can lack spatial resolution.(1) In built environments, reliable detection of SARS-CoV-2 has been shown with sampling of floors compared to other surfaces.(3,7) In long-term care homes, serial floor sampling may be particularly relevant as not all residents use toilets, and detection on floors was shown to predate COVID-19 outbreaks by days and even weeks.(19)

There have been numerous studies examining environmental presence of SARS-CoV-2 in school settings, however viral detection was infrequent, with test positivity ranging from 0% to 5%, despite the presence of student cases identified by clinical and asymptomatic screen testing.(5–7) As there was limited access to molecular testing in the community and lack of public reporting of cases, we examined publicly-reported absenteeism rates as a proxy measure for burden of illness among students and staff. However absenteeism reporting is highly variable across jurisdictions, with differences in ascertainment, reporting, and thresholds to trigger public health action.(20,21)

While environmental sampling of floors has been shown to correlate with COVID-19-related clinical activity in hospital and LTC settings, we did not find a correlation at the grade- or school-level. Also, PCR-based detection cannot discriminate among fresh or relic RNA, or RNA brought into buildings via fomites, and may not be useful for surveillance of active viral shedding at a single point in time.(7,22,23) Notably our outcomes were not true measures of COVID-19 burden within individual schools, and there may be value for establishing trends and following progression in test positivity as a reflection of community pediatric burden. There was clear heterogeneity between sites and age groups that may reflect immunity through vaccination and/or prior exposure to SARS-CoV-2. The role of surveillance in the built environment warrants further study to determine its usefulness in informing public health action to reduce the burden of communicable diseases.

## Data Availability

All data is not available for sharing.

## Acknowledgements

The authors would like to thank leaders and staff at the participating schools and Ottawa Public Health for their support and commitment to the success of this study.

---

**Supplemental Figure 1:**
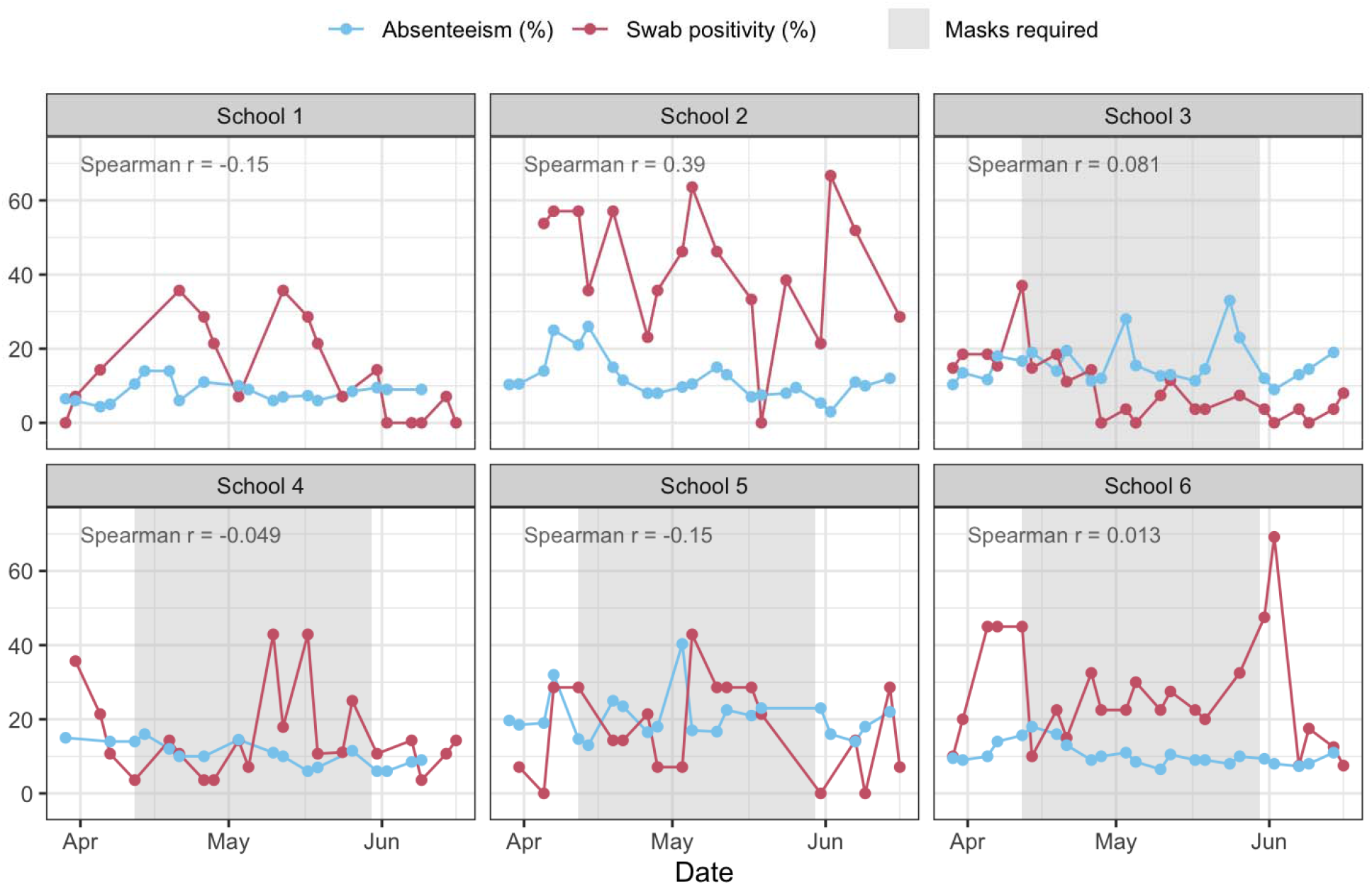
Time-series of biweekly aggregated absenteeism and floor swab positivity rates at the school level. Shaded areas indicate the period in which mandatory masking policies were reinstituted at the school level.

**Supplemental Figure 2:**
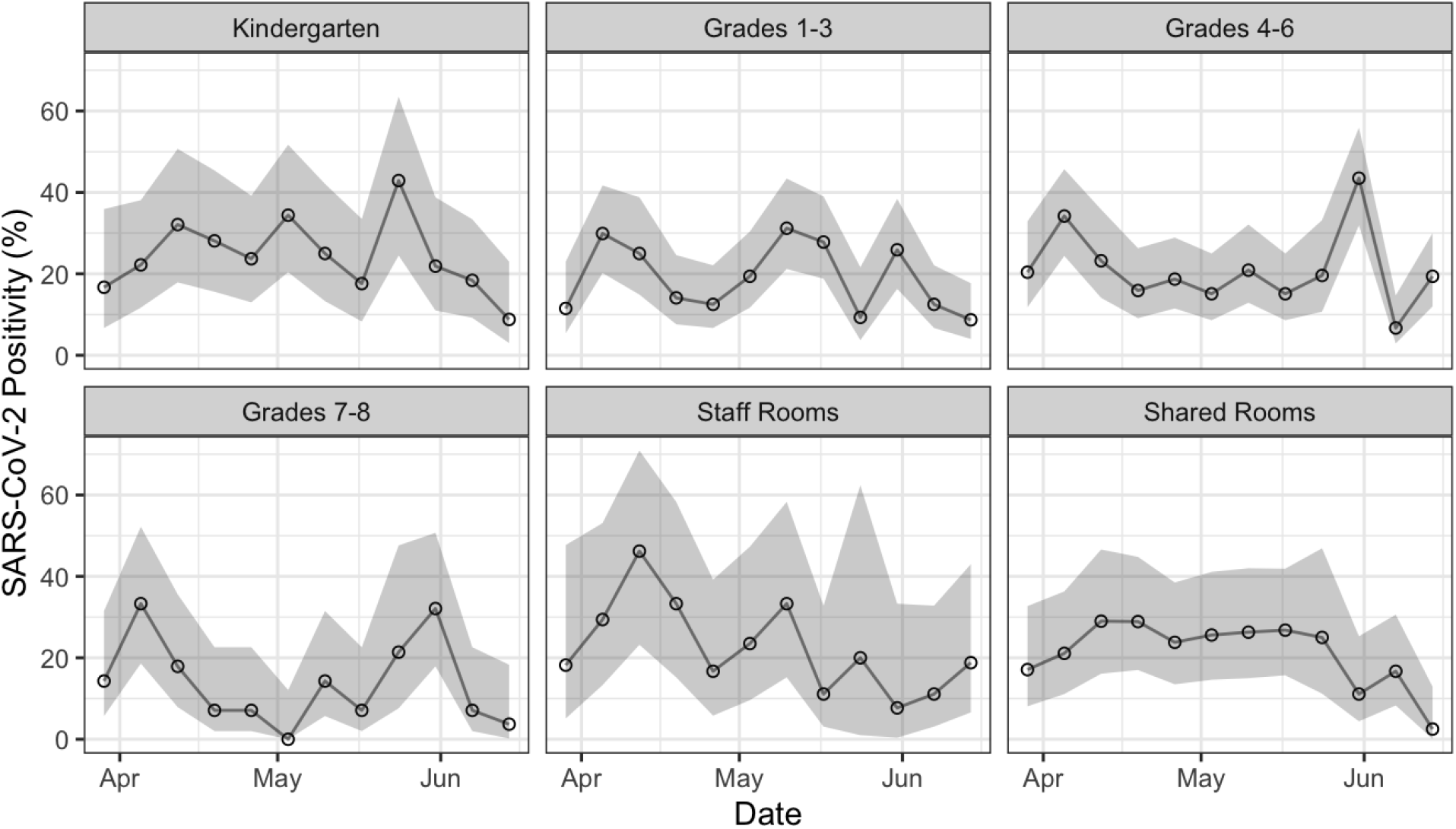
Weekly swab positivity aggregated by grade. **Shaded** areas represent 95% confidence intervals.

## REFERENCES

1. Manuel DG, Delatolla R, Fisman DN, Fuzzen M, Graber T, Katz GM, et al. The Role of Wastewater Testing for SARS-CoV-2 Surveillance [Internet]. Ontario COVID-19 Science Advisory Table; 2021 Aug [cited 2023 Jan 19]. Available from: https://covid19-sciencetable.ca/sciencebrief/the-role-of-wastewater-testing-for-sars-cov-2-surveillance

2. Fralick M, Nott C, Moggridge J, Castellani L, Raudanskis A, Guttman DS, et al. Detection of Covid-19 Outbreaks Using Built Environment Testing for SARS-CoV-2. NEJM Evidence [Internet]. 2023 Feb 28 [cited 2023 Mar 2];2(3). Available from: https://evidence.nejm.org/doi/10.1056/EVIDoa2200203

3. Hinz A, Xing L, Doukhanine E, Hug LA, Kassen R, Ormeci B, et al. SARS-CoV-2 detection from the built environment and wastewater and its use for hospital surveillance. Ivanova EP, editor. FACETS. 2022 Jan 1;7:82–97.

4. Ziegler MJ, Huang E, Bekele S, Reesey E, Tolomeo P, Loughrey S, et al. Spatial and temporal effects on severe acute respiratory coronavirus virus 2 (SARS-CoV-2) contamination of the healthcare environment. Infect Control Hosp Epidemiol. 2022 Dec;43(12):1773–8.

5. Crowe J, Schnaubelt AT, SchmidtBonne S, Angell K, Bai J, Eske T, et al. Assessment of a Program for SARS-CoV-2 Screening and Environmental Monitoring in an Urban Public School District. JAMA Netw Open. 2021 Sep 22;4(9):e2126447.

6. Cordery R, Reeves L, Zhou J, Rowan A, Watber P, Rosadas C, et al. Transmission of SARS-CoV-2 by children to contacts in schools and households: a prospective cohort and environmental sampling study in London. The Lancet Microbe. 2022 Nov;3(11):e814–23.

7. Zuniga-Montanez R, Coil DA, Eisen JA, Pechacek R, Guerrero RG, Kim M, et al. The challenge of SARS-CoV-2 environmental monitoring in schools using floors and portable HEPA filtration units: Fresh or relic RNA? Gregori L, editor. PLoS ONE. 2022 Apr 22;17(4):e0267212.

8. Manuel D, Yusuf W, Thomson M, Lee I. The Public Health Environmental Surveillance Database (PHESD) [Internet]. Zenodo; 2021 [cited 2023 Jan 19]. Available from: https://zenodo.org/record/5660201

9. Government of Ontario. 2022: School Absenteeism [Internet]. 2022 [cited 2023 Feb 19]. Available from: https://data.ontario.ca/dataset/summary-of-cases-in-schools/resource/e3214f57-9c24-4297-be27-a1809f9044ba

10. Ottawa Public Health. COVID-19 - COVID-19 in neighbourhoods from March 09 2020 to December 31 2021, accessed from Ottawa Neighbourhood Study/University of Ottawa [Internet]. 2022 [cited 2023 Feb 4]. Available from: https://www.neighbourhoodstudy.ca/covid-19-in-ottawa-neighbourhoods/

11. R Foundation for Statistical Computing. R: A language and environment for statistical computing. [Internet]. 2022. Available from: https://www.R-project.org/

12. Bates D, Mächler M, Bolker B, Walker S. Fitting Linear Mixed-Effects Models Using lme4. J Stat Soft [Internet]. 2015 [cited 2023 Feb 14];67(1). Available from: http://www.jstatsoft.org/v67/i01/

13. Andrejko KL, Head JR, Lewnard JA, Remais JV. Longitudinal social contacts among school-aged children during the COVID-19 pandemic: the Bay Area Contacts among Kids (BACK) study. BMC Infect Dis. 2022 Dec;22(1):242.

14. Hall CB, Douglas RG. Modes of transmission of respiratory syncytial virus. The Journal of Pediatrics. 1981 Jul;99(1):100–3.

15. Paul LA, Daneman N, Schwartz KL, Science M, Brown KA, Whelan M, et al. Association of Age and Pediatric Household Transmission of SARS-CoV-2 Infection. JAMA Pediatr. 2021 Nov 1;175(11):1151.

16. Lessler J, Grabowski MK, Grantz KH, Badillo-Goicoechea E, Metcalf CJE, Lupton-Smith C, et al. Household COVID-19 risk and in-person schooling. Science. 2021 Jun 4;372(6546):1092–7.

17. Lammie SL, Ford L, Swanson M, Guinn AS, Kamitani E, van Zyl A, et al. Test-to-Stay Implementation in 4 Pre–K-12 School Districts. Pediatrics. 2022 Oct 1;150(4):e2022057362.

18. World Health Organization. Environmental surveillance for SARS-COV-2 to complement public health surveillance [Internet]. 2022. Available from: https://www.who.int/publications/i/item/WHO-HEP-ECH-WSH-2022.1

19. Fralick M, Nott C, Moggridge J, Castellani L, Raudanskis A, Guttman DS, et al. Detection of COVID-19 Outbreaks in Long-Term Care Homes Using Built Environment Testing for SARS-CoV-2: A Multicentre Prospective Study [Internet]. Public and Global Health; 2022 Jun [cited 2023 Jan 19]. Available from: http://medrxiv.org/lookup/doi/10.1101/2022.06.28.22276560

20. Public Health Ontario. COVID-19: Use of Absenteeism Data to Inform Public Health Measures in K-12 Schools [Internet]. 2022. Available from: https://www.publichealthontario.ca/-/media/Documents/nCoV/sch/2022/02/es-absenteeism-data-ph-measures-K-12-schools.pdf?sc_lang=en

21. Egger JR, Hoen AG, Brownstein JS, Buckeridge DL, Olson DR, Konty KJ. Usefulness of School Absenteeism Data for Predicting Influenza Outbreaks, United States. Emerg Infect Dis. 2012 Aug;18(8):1375–7.

22. Krambrich J, Akaberi D, Ling J, Hoffman T, Svensson L, Hagbom M, et al. SARS-CoV-2 in hospital indoor environments is predominantly non-infectious. Virol J. 2021 Dec;18(1):109.

23. Zhou J, Otter JA, Price JR, Cimpeanu C, Meno Garcia D, Kinross J, et al. Investigating Severe Acute Respiratory Syndrome Coronavirus 2 (SARS-CoV-2) Surface and Air Contamination in an Acute Healthcare Setting During the Peak of the Coronavirus Disease 2019 (COVID-19) Pandemic in London. Clinical Infectious Diseases. 2021 Oct 5;73(7):e1870–7.

